# Perceptions of Water Security and Climate Change in Lima, Peru: Qualitative Study of Mothers and Healthcare Providers

**DOI:** 10.1101/2024.07.10.24309904

**Authors:** Danna Obregon Morales, Karen Ramos, Elena Jauregui, Milagros Dueñas, Nancy Rumaldo, Shruti Gogia, Leonid Lecca, Sonya S. Shin

## Abstract

**Background:** Water insecurity, a global public health crisis, will be intensified by climate change. In coastal Peru, little is known about the health effects of water insecurity from a community perspective. Understanding first-hand experiences and perceptions of mothers and healthcare providers can inform strategies to mitigate the effects of water insecurity and climate change on health outcomes and health disparities.

**Methods:** This qualitative study took place in 2023 in Carabayllo, the Northernmost region of Lima, Peru comprised of urban and rural communities. We conducted six focus groups with 10 pregnant women and 23 mothers of children ages 0 to 10, as well as individual interviews with 22 healthcare providers. Data were analyzed through rapid qualitative analysis combining deductive and inductive methods.

**Principal findings:** Water insecurity was common among community members and was normalized as part of daily life. Access to water varied based on socioeconomic status and geography. Perceived health impacts were primarily related to hygiene and sanitation (e.g. diarrheal disease), water storage (dengue), and longer-term effects such as nutrition and child development. Respondents agreed that climate change would exacerbate water insecurity and endorsed strategies to take action.

**Conclusions and significance:** The lived experiences of water insecurity and its health impacts in Lima, Peru highlight the importance of mobilizing community stakeholders, health systems and local government to increase water security and support healthcare facilities and communities on water conservation and climate preparedness efforts.

## Introduction

Approximately 2.2 billion people worldwide lack access to safely managed drinking water, with a significant concentration of this burden in LMICs (1). Climate change has exacerbated this crisis, leading to increased water scarcity, irregular precipitation patterns, and extreme weather events (2). The United Nations predicts climate change will further intensify water insecurity, affecting up to 3.2 billion people by 2050 (3).

In Peru, only 50% of the population has access to safe water (4). Drinking water availability varies across geographic regions: 92% of urban populations have access to potable water, compared with 79% of residents in rural areas (5). In terms of health consequences, mortality from inadequate water and sanitation services is disproportionately higher in the jungles and highlands compared to coastal regions; on the other hand, the dry coastal regions are the most vulnerable to climate-related water scarcity and immigration (6). According to the World Bank, despite ample renewable water resources, Peru’s water stress is largely driven by inadequate water infrastructure, population growth, and excessive water use by agricultural and industrial (e.g. manufacturing, mining) sectors (6).

Whereas lived experiences and perceptions among community members have been explored in other regions of Peru (7, 8), there is a paucity of individual experiential and perceptual data among urban, coastal Peruvian communities with respect to water security and climate change (9, 10). Understanding community members’ lived experiences with water security and awareness of climate change implications could help to assess community readiness and inform policy decisions and community-led strategies to mitigate future health effects of climate change, particularly related to water scarcity.

To address this gap, we conducted a qualitative study to explore community perspectives of water insecurity and its impact on maternal child health in Carabayllo, the Northernmost region of Lima, Peru. We sought to understand direct experiences of water insecurity, as well the perceived relationship of water insecurity to climate change and health outcomes from the perspectives of community members (mothers and pregnant women) as well as healthcare providers working in the region. In addition, we sought to identify existing strategies to mitigate the effects of climate change and water insecurity, implemented at household, community and governmental levels.

## Materials and Methods

### Study design and participants

This study was nested within a broader community needs assessment to characterize the supply and demand of healthcare services among pregnant women and children ages 0 to 10 in Carabayllo, Lima. The needs assessment was designed and carried out by Socios En Salud, a non-governmental organization operating in Carabayllo since 1995, to inform future programming. SES operates numerous community-based initiatives, many of which rely on community health workers (CHWs) and mobile technology to provide accompaniment support for various health conditions including maternal health, early child development and nutrition (11–13). Recognizing the intersection of climate health and water insecurity with these conditions, as well as the lack of relevant data pertaining to this population, we embedded additional questions into the needs assessment.

The study population included healthcare providers, pregnant women, and caregivers of children aged 0 to 10. Four healthcare establishments were selected among a total of 12 health establishments in Carabayllo to represent different facility types and populations served: one health post in a rural community, two health posts serving urban regions, and a health center in an urban neighborhood (14). For the needs assessment, pregnant women and children receiving care at these establishments were identified through chart review to generate a list of individuals who met eligibility criteria (pregnant woman or child aged 0 to 10). Pregnant women and caregivers of children were recruited to take part through community outreach, in which CHWs invited candidates to take part in the needs assessment, which involved a survey followed by focus groups with a subset of survey respondents.

Among individuals who completed the community survey, those who actively engaged in the survey (e.g. provided longer responses to open-ended questions, expressed enthusiasm for taking part in the survey) were consecutively invited to participate in focus groups. Those who expressed interest were provided the time and place of the group interview. A total of 10 participants were invited to each group interview (60 recruited in total), and the interview took place regardless of how many people showed up.

For healthcare providers, we conducted one-on-one interviews and used purposive sampling for diverse professional representation at each facility. We recruited the main provider caring for pregnant women and children in each of the following professions: nutritionist, obstetrician, head physician, odontologist, psychologist and nurse. Of note, in smaller facilities such as health posts, not all professions were present. In addition, three healthcare providers (two obstetricians and one nurse) working at the Ministry of Health District level were interviewed.

### Study setting

Located in the northernmost region of Lima, Carabayllo comprises 347 square kilometers, and has a population of 333,039 people (15). Given its location on the periphery of the capital city, Carabayllo represents a mix of urban and rural areas, including expanding neighborhoods fed by a continuous influx of immigrants from within and outside of the country. As Carabayllo has expanded, newer settlements (“asentamientos humanos” or if settled without permission “invasiones”) are typically located on steep, rocky hillsides (“cerros”) with minimal vegetation or infrastructure. As settlements gain formal recognition, they also obtain rights to municipal infrastructure such as water and electricity. More established communities (“urbanizaciones”) are typically more developed, with paved roads, local businesses such as shops and markets, and housing infrastructure such as paved floors, concrete construction, electricity and plumbing. Where services are available in Lima, the water utility company (Sedapal) provides potable water for a cost (16). The monthly household cost for water ranges from 10 to 300 ($3-79 USD), averaging 45 soles or approximately $12 USD (internal data).

### Data collection

The study team developed the following interview questions to explore experiences of water shortages, understandings of climate change and its relationship to water access, impacts of water insecurity and/or climate change on maternal child health, and practices to adapt to water scarcity in the face of climate change: *Have you ever had a time when your [family / community] did not have enough water to drink and use? [probe: if yes, describe]; What do you understand when you hear the words climate change? [if not spontaneously mentioned, probe perceived relation to water access]; Climate change can sometimes cause water problems including damage from flooding or water shortages due drought and heat. Do you feel that your [family / community] has been affected by climate change? [probe: if yes, describe]; Regardless of climate change, we know that Peru’s communities are resilient and have survived periods of drought over centuries. Does your [family / community] have any practices to conserve water or manage during times of water scarcity? [probe: if yes, describe].* For women and caregivers, we used the term “your family,” whereas for healthcare providers, we used “your community,” referring to residents of the catchment served.

A trained Peruvian study staff (DO) conducted all interviews in Spanish; this individual had no prior relationships with study participants. Interviews took place in-person at the health centers and community centers (“casas comunitarias de salud”) and were audio-recorded. We used rapid qualitative methods to analyze the interviews using a deductive-inductive approach. Rapid qualitative methods were utilized to complete the analysis within the study time-frame available and small research team. Two coders – one Native Spanish speaker (EJ) and one non-Native Spanish speaker (SSS) – established a draft codebook based on *a priori* domains (e.g. first-hand experience of water scarcity; perceived relationship of climate change with water shortages; health effects of water scarcity). Each coder then reviewed audio-recordings of 3-4 interviews, using the draft codebook to identify themes within each domain, tagging relevant citations using audio-recording timestamps. The interviewer and coders then met to review and refine the codebook to incorporate themes using an inductive approach. For example, under the domain “first-hand experience of water stress/scarcity,” the following themes were identified: variability in water supply based on geographic region, changes in water supply over time, interruptions/unreliability in water supply, diminished water security due to heat. The final codebook (**Table 1**) was then used by a single coder (SSS) to analyze all interviews, flagging all coded excerpts with timestamps. Coded excerpts were then translated and anonymized. All translations were reviewed and edited by a Native Spanish speaker (DO, EJ). While sampling was determined by funding rather than saturation, the analysis team did feel that we reached saturation among both participant groups.

**Table 1:**
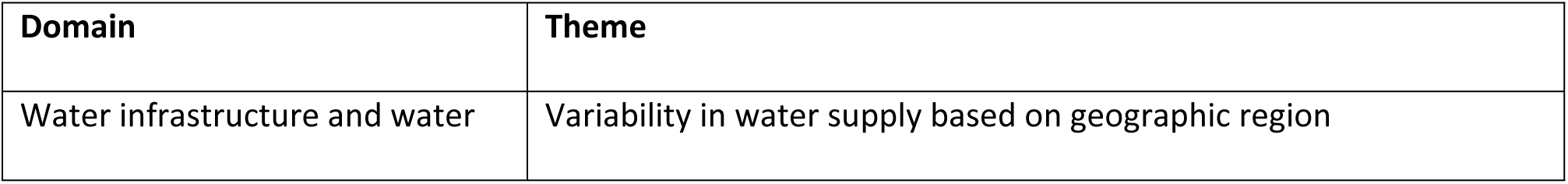
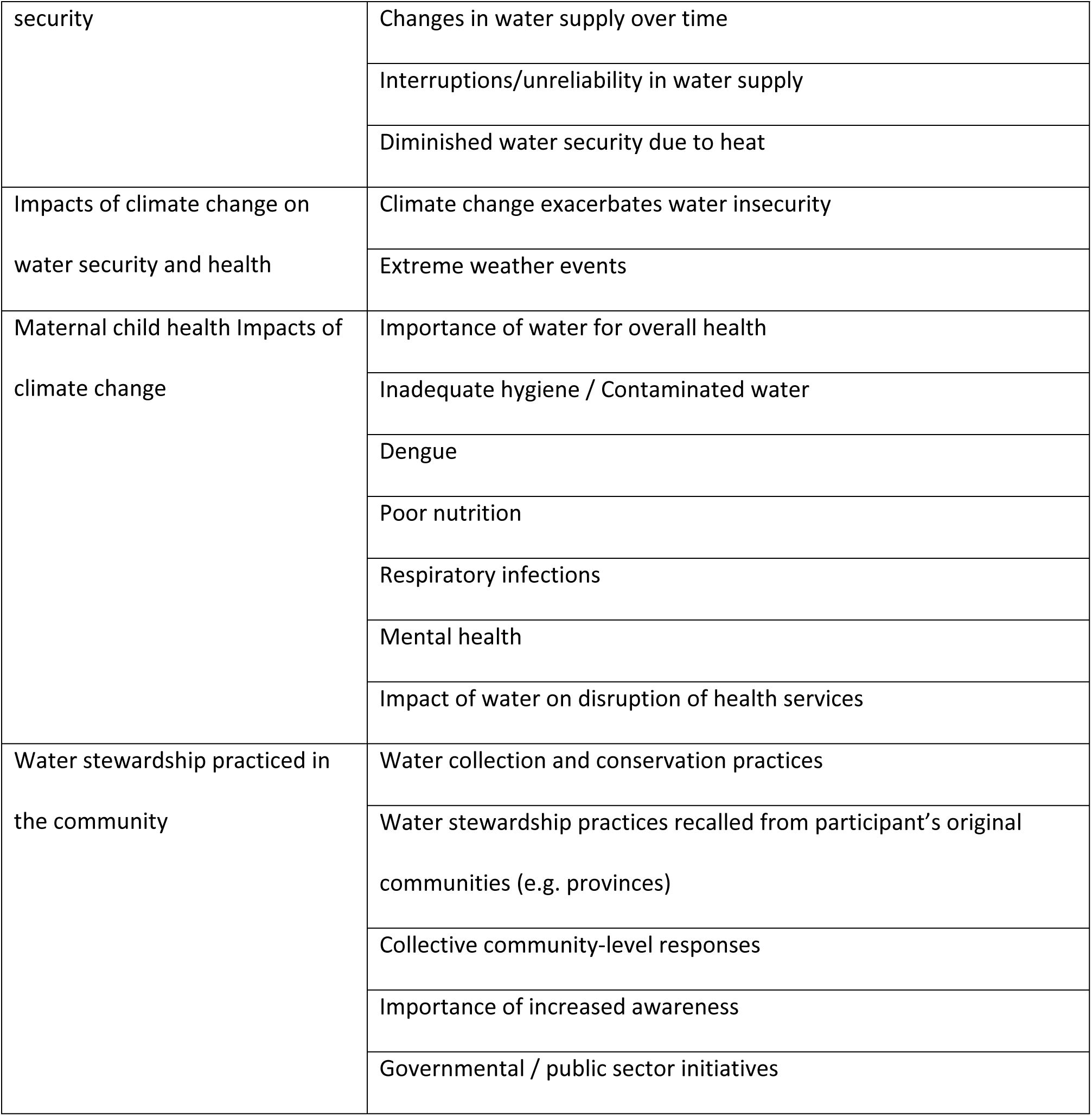
Domains and themes.

The core team (EJ, SSS, DO) met frequently throughout the analysis phase to confirm consistency and coherence of domains and themes, collectively identify key findings, select illustrative quotes, and relate themes into an overall schema. Preliminary findings including the overall schema were shared with the broader study team (DO, KR, EJ, MD, NR, LL, SG, JSP) for further feedback, interpretation and reflections.

### Ethical considerations and positionality

Study participants provided written informed consent for data collection and received a healthy meal as for compensation for their time. The cost of transportation to attend focus groups was also covered. Ethics committee approvals were obtained from Mass General Brigham (2023P002227) and Vía libre (022-2023) Institutional Review Boards. The study team – five Peruvian staff of Socios En Salud, two Public Health Masters students (one Peruvian, one non-Peruvian) and a non-Peruvian clinician-researcher with 29 years of experience working in Peru – carried out the study including study design (KR, MD, NR, SS, LL), data collection (DO), analysis (EJ, DO, SS), interpretation of study findings (DO, KR, EJ, MD, NR, SG, LL, SS, JSP), manuscript preparation (SG, SS, DO) and manuscript review (all authors).

## RESULTS

From August 22 through September 27, 2023, we conducted six focus groups and 26 individual interviews. Ten pregnant women took part in three focus groups and 23 caregivers of children aged zero to ten years old participated in three focus groups (Table 2). The median ages among pregnant women and caregivers were 28.5 years (interquartile range 23-34) and 32 (interquartile range 27-38), respectively. Although male caregivers were eligible to take part, all caregivers were mothers; for this reason, herein, we refer to the cohort of pregnant women and caregivers as “mothers.” In addition, 22 healthcare providers (four nurses, four physicians, three nutritionists, six obstetricians, four odontologists and one psychologist) were interviewed.

**Table 2:**
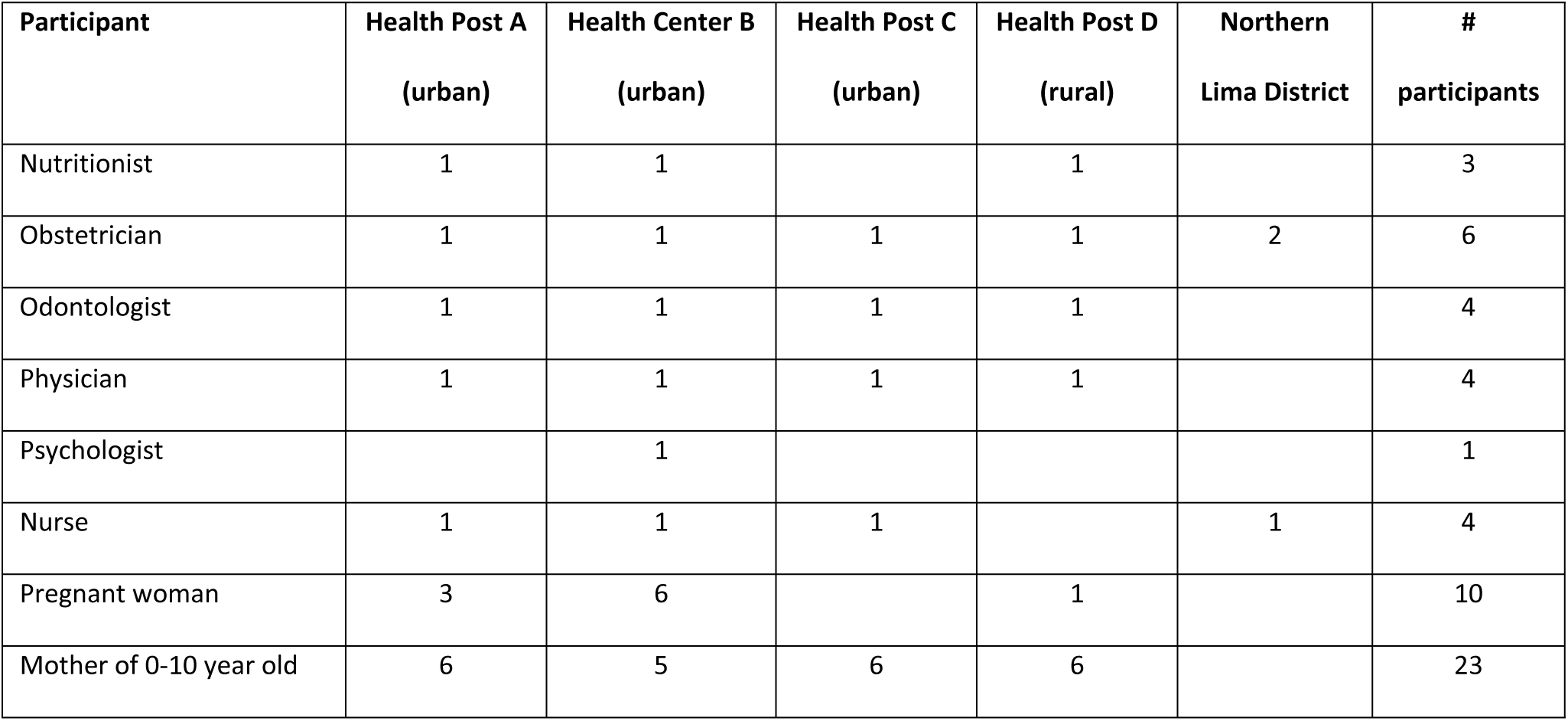
Participant summary.

### Water infrastructure and water security

Most communities described challenges with water, which varied based on geography and water source. More established zones (“urbanizaciones”) tended to have constant water supply, whereas less developed zones, such “asentamientos humanos” located on the hills (“cerros”) of Lima’s periphery, were more susceptible to interruptions in municipal water supply and unreliable water pressure when filling community cisterns or tanks. Other sites lacked any water infrastructure, and families had to pay for water delivered by truck. Another challenge for informal communities (“invasiones”) was the lack of governmental recognition and planning to have formal water systems installed by Sedapal. The degree of water access also reflected socioeconomic disparities, with more affluent communities and households tending to have more reliable access.

> *Yes, we’ve had problems, mostly because I live way up (“arriba”), we have always suffered for water, because the supply from cisterns don’t last. They [water trucks] only come every two weeks or at the end of the month. And if maybe you aren’t there and you miss a filling, they don’t fill your tank and then you’re stuck because you have to really measure your water. And if you run out in those 15 days, who’s going to give you water? Nobody. So, I’ve suffered this. Once I went for a month without water. I have a brother-in-law who lives close to the wall. I had to carry water from down there to way up high in buckets. -* Mother of 6 to 10 year-old seen at health post (FGM3 #4)

> *In the higher zones, [residents] don’t have access to water. They have to buy water or use a cistern [transported by truck] which has to travel up the hill, up to a certain point, because even beyond, there are dirt hills where cisterns can’t reach. -* Obstetrician at urban health post (OB4)

While providers and community members generally felt that water supply had improved over time, communities in less developed regions did not have consistent access to running water. Households only had running water during certain times of day and would typically store water in containers for use throughout the rest of the day. Interruptions in water service could occur without warning and remain for unpredictable lengths of time. Periods of heat also affected water availability, causing dry wells or lower water levels in cisterns. Experiences of water scarcity were often normalized such that respondents simply described water scarcity or unreliability as a part of their routine lives.

> *In the past, those things [water shortages] happened, but it would start in the middle of the night, they would shut off the water so that there’d be water during the day. From this time to that time of day in this district, there’s water; in another district, no… that’s how they started to ration water, but it’s not that we don’t have water all day. In the past, yes, there was water scarcity, but now not so much. -* Obstetrician at Northern Lima District (OB2)

> *Unexpectedly, sometimes you go to the market, then you get back to your house and there’s no water… You don’t know exactly [sometimes] there’s water all day, [other times] we wait for water to come back… two hours, three hours, sometimes it returns the following day. -* Mother of 0 to 5 year-old seen at health post (FGM1 #5)

> *We don’t have running water, so we use a well (“pozo”). When it’s hot, the well doesn’t have water… January, February [summer months in Peru]. They have to bring a pump and try to pump up more water. This goes on for several months, the hottest months. And then we have to go down to collect water because there isn’t enough pressure to get the water up to our homes. -* Pregnant woman seen at health center (FGG1 #3)

### Perceived relationship of climate change and water security

While a few respondents did not perceive current impacts of climate change on water-related health issues, most noted connections. When asked how climate change related to water scarcity, healthcare providers and community members often referred to rain shortages, increased temperatures, and drought occurring in Lima and other regions of Peru. Respondents noted that these manifestations of climate change caused environmental and agricultural problems, such as contaminated rivers and lack of water for irrigation, with subsequent effects on the availability and cost of food and ultimately on health. Several respondents speculated that climate change effects were likely to increase over time.

> *I’ve heard that’s what they say, that climate change may cause scarcity of water. Especially in zones of poverty, it’ll affect us even more. But I haven’t witnessed first-hand experiences of climate change in this community [served by health establishment].* - Physician at urban health post (MED2)

> *Obviously yes, climate change is very important; better put, yes, [climate change] can affect water supply. Here we live in Lima, no, because we have water that is always available. But in the provinces, they depend on water for their produce. Depending on the climate and weather, there’s quite a bit of water scarcity. -* Pregnant woman seen at health center (FGG2)

Providers and community members also perceived that climate change was associated with more frequent extreme precipitation events in Lima, resulting in flooding and mudslides. These events occasionally caused local destruction of homes and barriers to accessing healthcare, especially for pregnant women. A few respondents noted the effects of extreme weather on crops, resulting in higher cost and reduced availability of food.

> *When it rains too much, the mud starts to come and can take the houses. The mud is a problem, when it rains all night, where we live there, it is just mud. -* Mother of 6 to 10 year-old seen at health post (FGM3 #7)

> *From the phenomenon of El Niño, yes, there were rains, and houses in many zones in Northern Lima (“Lima Norte”) were affected… In the case of [name of neighborhood] it was along the Rimac River, generally because [the river] rises and that’s when houses start to get flooded. Especially if your house isn’t concrete, definitely the rain’s going to get in and in the end, it can destroy your house, or from the mudslides (“huaicos”) that happened. -* Obstetrician at Northern Lima District (OB1)

### Impacts of water insecurity and climate change on maternal child health

Respondents viewed water as essential for health and wellbeing. Providers and community members identified multiple health conditions linked to water scarcity and poor water quality. Providers cited hygiene as a key factor, including one’s ability to wash hands and bathe, clean the house, and prepare food safely (e.g. washing produce, cleaning dishes). Mothers facing water scarcity prioritized water for drinking and cooking over use for bathing and household chores. These constraints in hygiene practices were linked with dermatologic and gastrointestinal problems, especially among children.

> *Water is life, without it we can’t do anything - we can’t eat, washing, washing our hands. Since the pandemic, we’ve learned the importance of washing hands, cleanliness (“aseo”), food. Without water, we can be exposed to all types of illnesses. -* Pregnant woman seen at health center (FGG1 #2)

> *Non-potable water contributes to parasites which can affect pregnant women. Other forms of gastroenteritis and other illnesses can affect pregnant women, affects their nutrition, anemia, and the health of their baby. -* Nutritionist at health center (NUT2)

Both providers and community members also linked respiratory problems with climate change (due to exposure to temperature fluctuations and extremes) and water insecurity (limiting one’s ability to clean resulting in increased pathogens and indoor allergens).

> *Also, the environment [is affected by water scarcity]: without water, you can’t clean and there’s going to be more dust which will cause respiratory problems especially with children who are more allergic. Also, poor living conditions which could be conducive to a rise in tuberculosis. -* Physician in urban health post (MED2)

> *Climate change causes cough, colds, from the humidity. Especially the children, because they don’t wash their hands, and the humidity affects their cough, cold, especially because they can get sick more rapidly. If they are outside, then by the night they could be coughing because their defenses are weaker. I have to take all my children to school, even my baby, because there’s nobody to take care of them. -* Mother of 6-10 year-old seen at health post (FGM3)

> *For pregnant women, the main thing I’ve seen is respiratory illness. More because of the cold, they close up in their houses, and there’s less ventilation. -* Nutritionist at urban health post (NUT1)

Both providers and mothers associated the recent surge in dengue with weather changes. The combined effect of higher temperatures and water scarcity has resulted in routine storage of water in unsealed containers. At the same time, heavy rain leaves stagnant collections of water. Most mothers were aware of practices (e.g. covering stored water) to prevent dengue, but even so sometimes observed larvae in their stored water. Given limited access, families still used contaminated water, although not for consumption. Providers described increased cases and noted particular concern regarding dengue among pregnant women.

> *Yes [lack of water] affects [patient health] because if we do a home visit, there are illnesses like dengue because they deposit their water in their containers (“bidon”) or in open buckets. Then there are lot of illnesses, because you go to the homes and you’ll see that there’s no place to throw out water because there’s no plumbing, or maybe there’s stagnant water (“agua detenida”), or maybe their container doesn’t have a cover. -* Obstetrician at health center (OB3)

> *If you don’t cover the water, the water gets worms, and then you can’t drink it, it’s only good for washing. That happened to me, my husband left [the water container] open, and I found little worms, like little fish. -* Mother of 6-10 year old seen at health post (FGM3)

> *Now we have a sealed tank so that mosquitos can’t enter. We used to have covered buckets (“baldes”) but even so, mosquitos contaminated the water. -* Pregnant woman seen at health center (FGG1)

Nutritional deficiencies were also described by providers and mothers, including anemia, especially among children. Dietary deficiencies were associated with water scarcity and climate change, both through malabsorption from parasitosis and rising food cost due to declining agricultural production.

> *For children, the lack of water could cause problems with malnutrition, and potentially development, growth curve. We have seen a lot of children with anemia, similar to national rates. -* Nutritionist at rural health post (NUT3)

> *In the zones that were heavily affected by rains and mudslides (“hauicos”), there has been a decrease in production and an increase in prices. This affects not just the zone where the food is produced, but also other areas… I’ve noticed the prices have risen too much, like the cost of onions and limes. -* Pregnant woman seen at health center (FGG1)

> *For pregnant women, they require a balanced nutrition, and this is a zone with high need. They need quality nutrition. If you don’t have water, you can’t meet your basic nutritional or sanitary needs. In terms of nutrition, when there is no water, there is a lack of food production… Also, when there are floods, this disrupts crops. This results in higher prices and less supply and makes it harder for pregnant women to buy the foods to meet their nutritional need. -* Obstetrician at urban health post (OB4)

Several providers noted that extreme weather and flooding disrupted access to healthcare services. For instance, one provider described a pregnant woman who delivered in her home because she was unable to travel to the health center due to rain and mud. Some providers also speculated that health systems could face greater strains due to climate change, for which they felt their systems to be ill-prepared.

> *There were mudslides, and a delivery happened in the home because [the pregnant woman] wasn’t able to make it to the center. We had to make a home visit, and it was pure mud. We were barely able to make it to her home, just imagine what it would be like for a pregnant woman to get to the center. She could have come to the center earlier, but she waited and didn’t come in for care. We went to her on a home visit to help her get care at the Maternity Center. There’s a lack of access for us to get to the homes or vice versa. -* Obstetrician at urban health post (OB4)

> *In this zone, there isn’t water: the water cuts off and comes only at certain hours. But we have a tank [in the health post] so the tank is filled and supplies us with water here. The problem comes when there’s not enough water pressure to fill the tank above, or on the other hand, there’s been 24 hours when there wasn’t water. So, then there’s no water. But it’s very few times that that’s happened. I can’t see patients, the only thing I can do is offer palliative treatments since the machines operate on electricity and water. -* Odontologist at urban health post (OD3)

### Community responses to water scarcity and climate change

When asked about water conservation measures, few participants could describe any activity that they currently practiced. Some women who had migrated from rural provinces described practices from their original communities, including collection of rain and surface water. However, since these sources of water were not available in Lima, these practices were no longer applicable. Similarly, at the community level, respondents were not aware of water conservation practices. Women stated they needed more resources or technology in order to adopt more conservation measures. For example, while one woman stated she was able to purchase a 2000-liter water tank for her home, other women were unable to afford such items. Finally, one woman described informal networks of support in her neighborhood to give water to each other when facing scarcity.

> *I remember my grandmother… when it rained, since she had a thatched roof, she would put these large plastic containers (“tinas grandes, latones de plástico”). When it rained in that place, she’d let them fill up, let them sit out all night so the dirt settles. From there, she’d take only from the top, the water that was clean to use, the rest she’d throw out. That was when there were heavy rains, with thunder and everything. -* Mother of 0-5 year-old seen at health center (FGM2 #4)

> *The rainwater is stored, it is clear in the mountains. But here it’s dirty. The air (“medioambiente”) here is polluted. People burn trash, the cars and traffic… the air is polluted and there’s not a lot of rain. You can’t compare it to the jungle. The quality of life, in terms of environment, is better in the jungle. The air there is pure. -* Mother of 0-5 year-old seen at health post (FGM3)

> *Some people are resourceful about this [conserving water], but they should have more resources and technology. -* Pregnant women seen at health center (FGG2 #1)

> *There are a lot of people who don’t have [water]. They come and ask me for water. Since I rent, I have running water, so I would give them water. And sometimes I’d run out and then have to ask them for water. But since I gave them water, they’d give me water too. -* Mother of 0-5 year-old seen at health post (FGM3 #5)

Healthcare providers acknowledged the need for increased awareness and communications related to climate change. This included both communications and preparedness at healthcare facilities and mass outreach to the community at large. Several District-level providers were aware of governmental actions, including a cross-sector Ministry program to monitor water quality, plans to install underground cisterns for water storage, and programs to treat community water tanks with larvicides. By and large, however, local healthcare providers viewed climate change as outside their clinical purview and were unable to identify any type of coordinated cross-sector governmental or community response.

> *Climate change has a major role in the wellbeing and health of people. We are not prepared for what could come. We know about El Niño and mudslides (“huaicos”), we are aware of these, but there are deficiencies. We need more education and health promotion so that we are prepared to make changes. Getting prepared for extreme temperatures, especially for vulnerable populations, elders, pregnant women, people who are immunocompromised. I do think we should look at this and take up these activities of prevention and promotion to confront climate change in case it’s not something temporary. -* Obstetrician at urban health post (OB4)

## Discussion

This qualitative study conducted in Carabayllo, Lima, Peru, illuminates the intricate nexus between water insecurity, climate change, and maternal and child health (**Fig 1**). Lived experiences highlight the quotidian challenges posed by water scarcity, as well as the normalization of water insecurity as part of everyday life.

**Figure 1:**
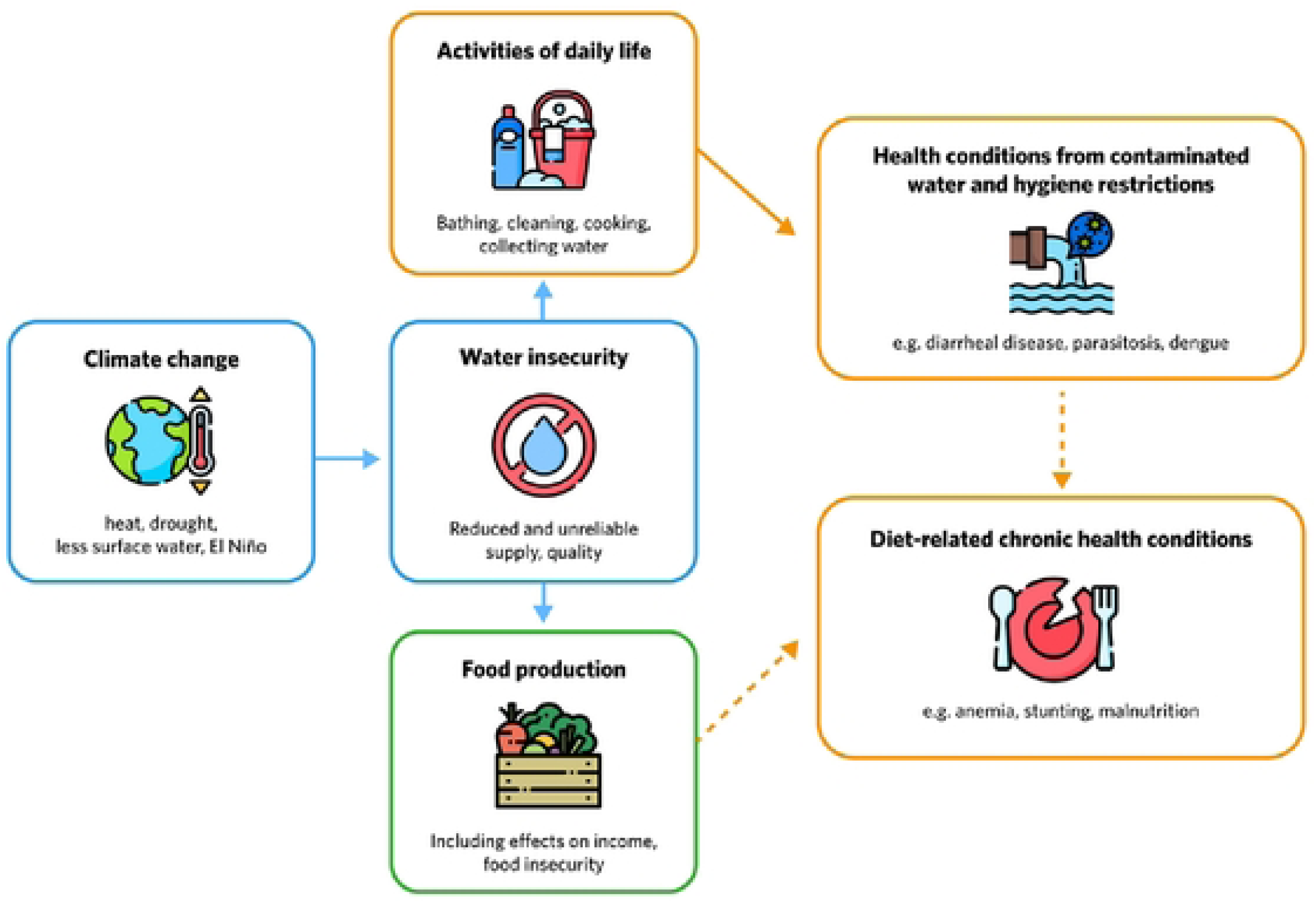
Inter-relationship between climate change, water insecurity, and maternal child health.

Our findings highlighted the repercussions of water scarcity on maternal and child health. Community members and healthcare providers observed acute health conditions (e.g. dermatological, gastrointestinal, respiratory) related to challenges in water, sanitation and hygiene, especially among children. In addition, longer-term effects of water scarcity on nutrition and child development were described. Effects of water insecurity on maternal and child health are well-described, reflecting the profound importance of access to clean water and sanitation on health and wellbeing (17–19). Water security is associated with improved child growth and development, as well as reduced under-five mortality; conversely, lack of potable water increases the risk of childhood diarrheal and dermatologic disease, stunting, maternal mortality, and vector-borne illnesses such as dengue (18, 20, 21). Furthermore, chronic under-hydration is associated with premature mortality and a spectrum of chronic diseases postulated to stem from accelerated aging (22).

We observed geographical and socio-economic disparities in water security, with more developed urban areas enjoying relatively constant supply compared to less developed zones which must grapple with interruptions and unreliable pressure. Socioeconomic disparities in water insecurity are well-documented and are likely to increase with climate change (23). Water insecurity is determined by a constellation of factors including water availability, quality, infrastructure, affordability and territoriality (24). Territoriality, which refers to the ability of local government to guarantee equitable access to water, requires both adequate water supply as well as political will for equitable distribution of resources (25). Prioritizing timely implementation of policies and strategies to ensure equitable access to safe drinking water is crucial, including infrastructure development directed toward the most vulnerable communities (4). In addition, identifying at-risk households and providing resources to increase water security and address underlying poverty could reduce health disparities (26).

In our study, healthcare providers and mothers had varying perceptions of climate change impacts. While some perceived climate change as imminent, others viewed it as a distant concern that was likely to increase over time. These findings are consistent with prior studies carried out in Peru. In 2021, Torres-Slimming et al. examined perceptions of climate change among teenagers and key stakeholders in three Peruvian cities (Cusco, Piura and Loreto) and found that community members were able to identify extreme weather events related to climate change, and in particular, raised concerns about water insecurity (9). Similarly, a mixed-methods study conducted in Cusco on community perceptions of climate change revealed that water scarcity emerged as one of the most prominent issues associated with climate change (7). In this 2021 study, some community members were able to describe adaptation strategies that could mitigate the long-term effects of climate change in their communities. In contrast, Sina et al interviewed Lima municipal officials in 2016 and reported limited awareness and understanding of climate change, along with low prioritization of climate change as a public health priority (10). Interestingly, a study of environmental practices among nurses in Northern coastal Peru found that most nurses practiced measures such as conserving energy use and recycling; the authors highlight the importance of engage healthcare providers in adopting and modeling environmental practices as trusted role models in the workplace and community (27).

Despite common water shortages, few healthcare providers or mothers were currently practicing water conservation measures. Healthcare providers expressed collective unpreparedness for impending impacts of climate change on healthcare services, signaling a gap in institutional readiness and the need for proactive response. We perceived community readiness, as providers and community members expressed a need to raise awareness, build water conservation skills, and provide additional resources to support healthcare facilities and communities to promote water conservation and climate preparedness. The World Health Organization provides guidance to assess and advance climate preparedness of healthcare facilities (28). Contingencies for transportation and community-based delivery of care – especially for high-risk groups such as women, children and elders – could further enhance the resiliency of healthcare systems (29, 30). At the community level, mothers expressed the need for more resources and technology to implement effective water conservation measures, citing examples of water storage equipment that would protect household vulnerability to water scarcity. The strength of existing informal support networks – such as sharing water on the principle of reciprocity – could serve as a foundation to explore community-based water co-operatives.

This study from Lima, Peru contributes to a growing body of research by providing context-specific insight into the diverse ways that water insecurity influences maternal and child health, as well as the potential repercussions of climate change on this dynamic. Community readiness was evident among mothers and healthcare providers, who supported raising community awareness, increasing water infrastructure and conservation practices, and preparing healthcare systems. Based on these findings, we describe potential strategies that could mitigate the negative impacts of climate change and water insecurity on health and wellbeing.

At the local governance level:

1. Provide subsidies on water storage and conservation tanks.
2. Prioritize investments in integrated water resource management.

At the health system level:

1. Train healthcare providers, administrators and community health workers to enhance knowledge of climate change and health impacts and their ability to identify and respond to local related health risks.
2. Assess climate preparedness of healthcare facilities and implement policies to increase resilience.
3. Develop surveillance capacity and early warning systems for climate-related health hazards e.g. dengue outbreaks.

At the community level:

1. Establish water cooperatives with infrastructure, financial and regulatory management carried out through community governance.
2. Promulgate knowledge-sharing and facilitate adoption of water conservation and safe storage.
3. Provide support to households experiencing water insecurity to facilitate access to healthcare and minimize water and food insecurity.

Further discussions with community stakeholders, municipal leaders, and cross-sector Ministry leadership would be needed to identify optimal and feasible strategies.

Our study has several limitations including a relatively small sample size, potentially limiting the generalizability of findings. Additionally, the qualitative nature of the study restricts our ability to confirm perceived relationships between water access and health outcomes. Despite these limitations, this study provides valuable insight, serving as a foundation for future quantitative research as well as intervention strategies to address the complex interplay of water scarcity, climate change, and maternal-child health.

## Conclusion

Families in Carabayllo, Lima, currently experience water insecurity and perceived adverse effects on health, which is likely to increase with climate change. Cross-sector stakeholder engagement should identify suitable adaptive strategies to mitigate the impacts of water scarcity and climate change on maternal and child health. Once identified, these strategies can be implemented and evaluated for impact. Collaborative efforts between health departments, local governments, and community stakeholders will be crucial for developing and implementing policies that enhance water infrastructure, promote sustainable practices, and build resilience in the face of climate-related challenges.

## Data Availability

Transcript excerpts will be available upon request.

## Acknowledgments

We wish to acknowledge the community members and healthcare providers of Carabayllo who generously shared their time and experiences. We also wish to acknowledge the generous gift from Ronda Stryker to the Harvard Medical School Department of Global Health and Social Medicine’s research core which made this work possible.

## References

1. Wolf J, Johnston RB, Ambelu A, Arnold BF, Bain R, Brauer M, et al. Burden of disease attributable to unsafe drinking water, sanitation, and hygiene in domestic settings: a global analysis for selected adverse health outcomes. Lancet. 2023;401(10393):2060–71.

2. Water – at the center of the climate crisis. United Nations; 2024.

3. United in Science report: Climate Change has not stopped for COVID19 [press release]. United Nations September 9, 2020 2020.

4. Hernandez-Vasquez A, Rojas-Roque C, Marques Sales D, Santero M, Bendezu-Quispe G, Barrientos-Gutierrez T, et al. Inequalities in access to safe drinking water in Peruvian households according to city size: an analysis from 2008 to 2018. Int J Equity Health. 2021;20(1):133.

5. (INEI) NIoSaI. Peru: Forms of access to water and basic sanitation. 2016.

6. Peru: Strategic Actions Toward Water Security. The World Bank Global Water Security and Sanitation Partnership; 2023.

7. Brügger A, Tobias R, Monge-Rodríguez FS. Public Perceptions of Climate Change in the Peruvian Andes. Sustainability. 2021;13(5).

8. Tallman PS, Collins SM, Chaparro MP, Salmon-Mulanovich G. Water insecurity, self-reported physical health, and objective measures of biological health in the Peruvian Amazon. Am J Hum Biol. 2022;34(12):e23805.

9. Torrres-Slimming PA, López Flórez L, Castañeda Checa K, Durand Galarza O, Tallman PS, Salmon-Mulanovich G. Explorando percepciones del impacto del cambio climático en tres regiones en el Perú Revista Kawsaypacha Sociedad y Medio Ambiente. 2021;8:101–17.

10. Sina M, Wood RC, Saldarriaga E, Lawler J, Zunt J, Garcia P, et al. Understanding Perceptions of Climate Change, Priorities, and Decision-Making among Municipalities in Lima, Peru to Better Inform Adaptation and Mitigation Planning. PLoS One. 2016;11(1):e0147201.

11. Tzelios C, Contreras C, Istenes B, Astupillo A, Lecca L, Ramos K, et al. Using digital chatbots to close gaps in healthcare access during the COVID-19 pandemic. Public Health Action. 2022;12(4):180–5.

12. Miller AC, Rumaldo N, Soplapuco G, Condeso A, Kammerer B, Lundy S, et al. Success at Scale: Outcomes of Community-Based Neurodevelopment Intervention (CASITA) for Children Ages 6-20 months With Risk of Delay in Lima, Peru. Child Dev. 2021;92(6):e1275–e89.

13. Contreras C, Rumaldo N, Lindeborg MM, Mendoza M, Chen DR, Saldana O, et al. Emotional Experiences of Mothers Living With HIV and the Quest for Emotional Recovery: A Qualitative Study in Lima, Peru. J Assoc Nurses AIDS Care. 2019;30(4):440–50.

14. Analisis de Situación de Salud 2022: Direccion de Redes Integradas de Salud Lima Norte Peru Ministerio de Salud 2022.

15. Estadistica Poblacional Peru Ministry of Health [Available from: https://www.minsa.gob.pe/reunis/data/poblacion_estimada.asp.

16. ASIGNACIONES MAXIMAS DE CONSUMO - POR AGUA Y ALCANTARILLADO - GRUPO TARIFARIO 1 DE 4: Sedapal; 2024 [Available from: https://cdn.www.gob.pe/uploads/document/file/4242604/3989288-asignaciones-maximas-de-consumo-2024.pdf?v=1705417674.

17. Miller JD, Workman CL, Panchang SV, Sneegas G, Adams EA, Young SL, et al. Water Security and Nutrition: Current Knowledge and Research Opportunities. Adv Nutr. 2021;12(6):2525–39.

18. Hutton G, Chase C. The Knowledge Base for Achieving the Sustainable Development Goal Targets on Water Supply, Sanitation and Hygiene. Int J Environ Res Public Health. 2016;13(6).

19. Schuster RC, Butler MS, Wutich A, Miller JD, Young SL, Household Water Insecurity Experiences-Research Coordination N. “If there is no water, we cannot feed our children”: The far-reaching consequences of water insecurity on infant feeding practices and infant health across 16 low- and middle-income countries. Am J Hum Biol. 2020;32(1):e23357.

20. Akanda AS, Johnson K. Growing water insecurity and dengue burden in the Americas. Lancet Planet Health. 2018;2(5):e190–e1.

21. Benova L, Cumming O, Campbell OM. Systematic review and meta-analysis: association between water and sanitation environment and maternal mortality. Trop Med Int Health. 2014;19(4):368–87.

22. Dmitrieva NI, Gagarin A, Liu D, Wu CO, Boehm M. Middle-age high normal serum sodium as a risk factor for accelerated biological aging, chronic diseases, and premature mortality. EBioMedicine. 2023;87:104404.

23. Young SL, Bethancourt HJ, Ritter ZR, Frongillo EA. Estimating national, demographic, and socioeconomic disparities in water insecurity experiences in low-income and middle-income countries in 2020-21: a cross-sectional, observational study using nationally representative survey data. Lancet Planet Health. 2022;6(11):e880–e91.

24. Compact UG. Secretariat CWM, editor2017. [May 9, 2024]. Available from: https://ceowatermandate.org/posts/water-scarcity-water-stress-water-risk-actually-mean/.

25. Urquiza A, Billi M. Documentos de Proyectos Seguridad hídrica y energética en América Latina y el Caribe: Definición y aproximación territorial para el análisis de brechas y riesgos de la población. United Nations CEPAL Cooperacion Alemana; 2020.

26. Stoler J, Pearson AL, Staddon C, Wutich A, Mack E, Brewis A, et al. Cash water expenditures are associated with household water insecurity, food insecurity, and perceived stress in study sites across 20 low- and middle-income countries. Sci Total Environ. 2020;716:135881.

27. Rojas-Perez HL, Diaz-Vasquez MA, Diaz-Manchay RJ, Zena-Nanez S, Failoc-Rojas VE, Smith D. Nurses’ Environmental Practices in Northern Peruvian Hospitals. Workplace Health Saf. 2024;72(2):68–74.

28. WHO GUIDANCE FOR CLIMATE RESILIENT AND ENVIRONMENTALLY SUSTAINABLE HEALTH CARE FACILITIES. Geneva: World Health Organization; 2020.

29. Palinkas LA, O’Donnell ML, Lau W, Wong M. Strategies for Delivering Mental Health Services in Response to Global Climate Change: A Narrative Review. Int J Environ Res Public Health. 2020;17(22).

30. Climate Change and the Health of Pregnant, Breastfeeding, and Postpartum Women: U.S. Environmental Protection Agency; [Available from: https://www.epa.gov/climateimpacts/climate-change-and-health-pregnant-breastfeeding-and-postpartum-women.

